# Convalescent COVID-19 patients are susceptible to endothelial dysfunction due to persistent immune activation

**DOI:** 10.1101/2020.11.16.20232835

**Authors:** Florence WJ Chioh, Siew-Wai Fong, Barnaby E. Young, Kan-Xing Wu, Anthony Siau, Shuba Krishnan, Yi-Hao Chan, Louis LY Teo, Fei Gao, Ru San Tan, Liang Zhong, Angela S. Koh, Seow-Yen Tan, Paul A. Tambyah, Laurent Renia, Lisa FP Ng, David C Lye, Christine Cheung

**Affiliations:** Lee Kong Chian School of Medicine, Nanyang Technological University, Singapore; Institute of Molecular and Cell Biology, Agency for Science, Technology and Research, Singapore; National Centre for Infectious Diseases, Singapore; Department of Infectious Diseases, Tan Tock Seng Hospital, Singapore; A*STAR ID Labs, Agency for Science, Technology and Research, Singapore; Singapore Immunology Network, Agency for Science, Technology and Research, Singapore; Yong Loo Lin School of Medicine, National University of Singapore, Singapore; National Heart Centre Singapore, Singapore; Duke-NUS Medical School, Singapore; Department of Biological Sciences, National University of Singapore, Singapore; Division of Clinical Microbiology, Department of Laboratory Medicine, Karolinska Institute, ANA Futura, Campus Flemingsberg, Stockholm, Sweden; Department of Biochemistry, Yong Loo Lin School of Medicine, National University of Singapore; Department of Infectious Diseases, Changi General Hospital, Singapore; Department of Medicine, National University Hospital, Singapore

## Abstract

The rapid rise of coronavirus disease 2019 patients who suffer from vascular events after their initial recovery is expected to lead to a worldwide shift in disease burden. We aim to investigate the impact of COVID-19 on the pathophysiological state of blood vessels in convalescent patients. Here, convalescent COVID-19 patients with or without preexisting conditions (i.e. hypertension, diabetes, hyperlipidemia) were compared to non-COVID-19 patients with matched cardiovascular risk factors or healthy participants. Convalescent patients had elevated circulating endothelial cells (CECs), and those with underlying cardiovascular risk had more pronounced endothelial activation hallmarks (ICAM1, P-selectin or CX3CL1) expressed by CECs. Multiplex microbead-based immunoassays revealed some levels of cytokine production sustained from acute infection to recovery phase. Several proinflammatory and activated T lymphocyte-associated cytokines correlated positively with CEC measures, implicating cytokine-driven endothelial dysfunction. Finally, the activation markers detected on CECs mapped to the counter receptors (i.e. *ITGAL, SELPLG*, and *CX3CR1*) found primarily on CD8+ T cells and natural killer cells, suggesting that activated endothelial cells could be targeted by cytotoxic effector cells. Clinical trials in preventive therapy for post-COVID-19 vascular complications may be needed.

**Graphical abstract:** 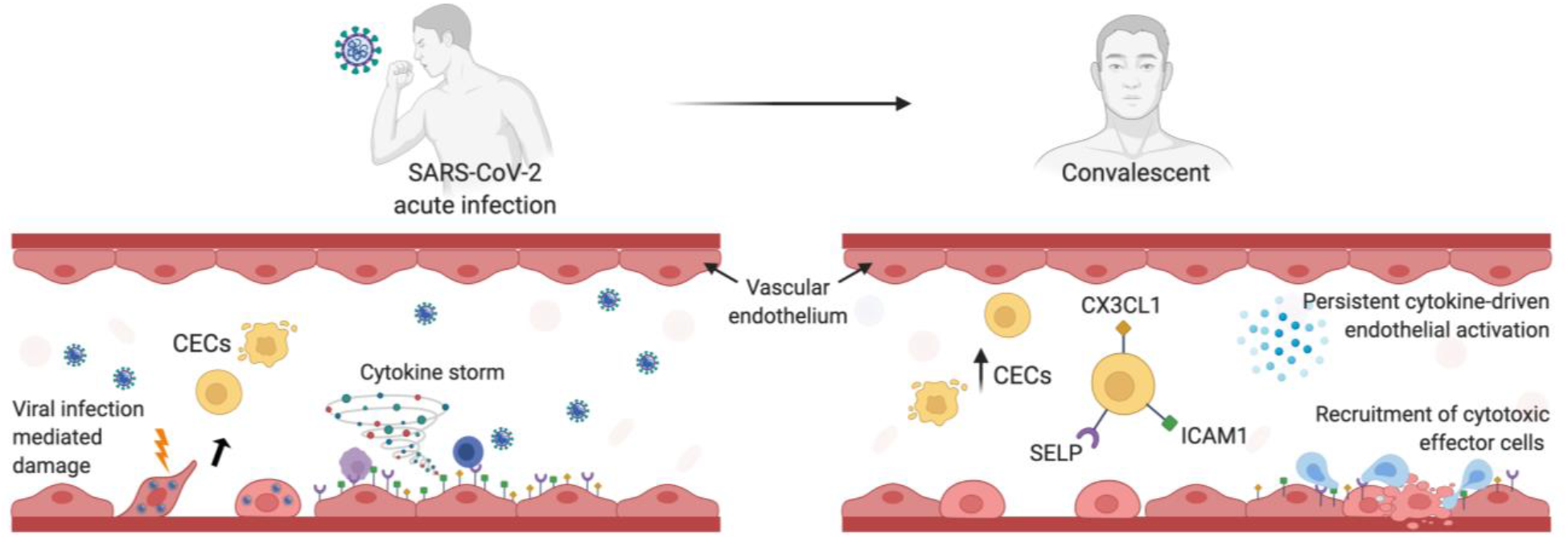

## INTRODUCTION

As of 11 October 2020, there have been more than 37 million confirmed cases and 1 million deaths from coronavirus disease 2019 (COVID-19). While many countries are still in their first wave of infection, some countries are now facing a second or third wave of resurgence upon reopening their economy, prompting new lockdown. Intensive ongoing research has shed light on the pathogenesis of COVID-19 and the extent of damages caused directly or indirectly by severe acute respiratory syndrome coronavirus 2 (SARS-CoV-2). Conversely, the intermediate and long-term complications of COVID-19 remains unclear (Del Rio, Collins et al., 2020). While most infected people recover completely within a few weeks, a considerable proportion continue to experience symptoms after their initial recovery (Yelin, Wirtheim et al., 2020) similar to SARS survivors (Ngai, Ko et al., 2010).

COVID-19 is mainly a respiratory infection. However, both autopsy findings and clinical observations have described vascular damages and thrombotic complications in a wide range of organs (Gupta, Madhavan et al., 2020) (Price, McCabe et al., 2020). Acute pulmonary edema was observed in severe COVID-19 patients with occlusion and thrombosis of pulmonary microvasculatures (Xu, Shi et al., 2020). A study of renal tissues found no vasculitis, hemorrhage or interstitial inflammation, but revealed endothelial activation and even frank necrosis (Su, Yang et al., 2020). The multi-organ impact of COVID-19 may increase the risk of long-term complications after recovery.

Importantly, systemic presence of the SARS-CoV-2 entry factor, angiotensin-converting enzyme 2 (ACE2) on the surface of endothelial cells, smooth muscle cells and pericytes suggests that the virus, once present in the circulation, may disseminate beyond the respiratory tract via the cardiovascular system (Hamming, Timens et al., 2004, Hoffmann, Kleine-Weber et al., 2020). While SARS-CoV is mainly a respiratory tract disease, the tropism of SARS-CoV-2 for vascular endothelial cells can be plausibly explained by the ability of SARS-CoV2 to use multiple viral entry-associated proteins expressed at various level in the vascular bed (Wazny, Siau et al., 2020). Underlying a viral-induced vascular dysfunction, multisystem inflammatory syndrome and chilblains reported in children and young adults are caused by inflammation of blood vessel walls and characterized by immune cell infiltration and thrombotic events (Colmenero, Santonja et al., 2020, Dolhnikoff, Ferreira Ferranti et al., 2020). Similarly, autopsy of COVID-19 patients’ tissues revealed evidence of endothelium damage resulting of direct viral infection together with extensive thrombosis, microangiopathy and T cell infiltration (Ackermann, Verleden et al., 2020, Varga, Flammer et al., 2020). Endothelialitis occurs in different organs as a consequence of viral infection and overactive host immune response.

Systematic evaluation of the long-term sequelae of COVID-19 is currently lacking. COVID-19 infection can cause extrapulmonary pathologies that persist after recovery such as cardiovascular events with ongoing myocardial inflammation (Puntmann, Carerj et al., 2020) acute ischemic or hemorrhagic stroke (Mao, Jin et al., 2020) as well as liver injury, neurological deficits, and acute kidney injury necessitating dialysis (Gupta et al., 2020). The thrombotic complications of COVID-19, such as pulmonary embolism may lead to lasting organ failure (Price et al., 2020). Independent of acute respiratory distress syndrome, severe pneumonia has been consistently associated with augmented risk of cardiac impairment both during convalescence and in later years (Corrales-Medina, Alvarez et al., 2015). We hypothesize that the risk of vascular complications in COVID-19 survivors will be higher, if compounded by a confluence of SARS-CoV-2-mediated damages, pro-inflammatory cytokine overdrive and comorbidities with hypertension and diabetes.

Circulating endothelial cells (CECs), dislodged from blood vessels as a consequence of vascular injury, constitute an ideal cell-based biomarker as it is indicative of *in situ* endothelial pathophysiology (Farinacci, Krahn et al., 2019, Hill, Zalos et al., 2003). Although CECs are a rare population in peripheral blood, they reflect endothelial dysfunction from a variety of vascular disorders including myocardial infarction, acute ischemic stroke, atherosclerosis and vasculitis (Nadar, Lip et al., 2005, Schmidt, Manca et al., 2015). In COVID-19, there are conflicting reports as to the frequency of CEC counts compared with healthy controls (Mancuso, Gidaro et al., 2020, Nizzoli, Merati et al., 2020). This discrepancy may reflect disease severity as COVID-19 patients admitted to the intensive care unit (ICU) have a higher CEC count, suggesting more pronounced endothelial injury in severe COVID-19 (Guervilly, Burtey et al., 2020). Interestingly, among non-ICU patients, those with chronic kidney disease had significantly higher CEC counts, implying that patients with pre-existing conditions may be more susceptible to vascular damage (Guervilly et al., 2020).

This study aims to understand the state of vascular health in convalescent COVID-19 patients, and to evaluate subclinical endothelial dysfunction through the phenotyping of CECs.

## RESULTS

### Patient and healthy participants characterization

To understand the intermediate consequence of COVID-19, we performed vascular phenotyping using CECs and endothelial activation markers as the cellular and molecular measures of endothelial dysfunction. Written informed consent was received from participants prior to inclusion in the PROTECT study (Young, Fong et al., 2020a, Young, Ong et al., 2020c). All study groups were almost gender-balanced, with prior comorbidities and self-identified ethnicity/race summarized in **Table 1**. Convalescent COVID-19 patients who had no pneumonia throughout admission (mild); pneumonia without hypoxia (moderate) or pneumonia with hypoxia (desaturation to ≤94%) requiring supplemental oxygen (severe), and discharged for 15 days [median (interquartile range, IQR 11-26)] were screened for pre-existing cardiovascular risk factors. Among our selected convalescent COVID-19 patients (n = 30), half had at least one cardiovascular risk factor including mainly hypertension, diabetes and/or hyperlipidemia. They were benchmarked to healthy participants (n = 13) and non-COVID patients with matched cardiovascular risk factors (n = 20). More details more demographics are included in **Table 1**.

**Table 1:** Demographics of patients and healthy controls.

| Characteristics, N (%) | Healthy participants<br>(n = 13) | Non-COVID-19 patients with cardiovascular risk factors<br>(n = 20) | Convalescent COVID-19 individuals<br>(n = 30) |  |
| --- | --- | --- | --- | --- |
|  |  |  | Cardiovascular risk factors (n=15) | No cardiovascular risk (n=15) |
| Age* | 40.3 (6.7) years | 62.4 (8.7) years | 54 (8.7) years | 42 (13.5) years |
| Age, Male* | 40 (15.2) years | 62.3 (10.4) years | 54 (8.2) years | 36 (6.1) years |
| Age, Female* | 40.7 (8.1) years | 63 (7.3) years | 54 (9.9) years | 48 (17.1) years |
| Gender, Male | 7 (53.8) | 10 (50) | 8 (53.3) | 8 (53.3) |
| Gender Female | 6 (46.2) | 10 (50) | 7 (46.7) | 7 (46.7) |
| Ethnicity | Chinese: 11 (84.6) | Chinese: 17 (85) | Chinese: 11 (73.3) | Chinese: 13 (86.6) |
|  | Filipino: 2 (15.4) | Indian: 3 (15) | Malay: 2 (13.3) | Indian: 1 (6.7) |
|  |  |  | Bangladeshi: 1 (6.7) | Caucasian: 1 (6.7) |
|  |  |  | Filipino: 1 (6.7) |  |
| Days post-symptom onset [median (IQR)] | N.A. | N.A. | 34 (27-46) days |  |
| COVID-19 severity | N.A. | N.A. | Mild: 5 (33.3) | Mild: 5 (33.3) |
|  |  |  | Moderate: 5 (33.3) | Moderate: 5 (33.3) |
|  |  |  | Severe: 5 (33.3) | Severe: 5 (33.3) |
| Comorbidities: |  |  |  |  |
| Hypertension | 0 (0) | 20 (100) | 10 (66.7) | 0 (0) |
| Hyperlipidemia | 0 (0) | 10 (50) | 11 (73.3) | 0 (0) |
| Diabetes mellitus | 0 (0) | 9 (45) | 7 (46.7) | 0 (0) |
| Fatty Liver | 0 (0) | Information not available | 2 (13.3) | 0 (0) |
| Chronic Liver Disease | 0 (0) | Information not available | 1 (6.7) | 0 (0) |
| Coronary Artery Disease | 0 (0) | 6 (30) | Information not available | Information not available |
| Myocardial infarction | 0 (0) | 0 (0) | 2 (13.3) | 0 (0) |

All values are reported as N (%) where N indicted number of observations. \*Values are expressed as mean ( ± standard deviation).

### COVID-19 and cardiovascular risk factors contribute to endothelial dysfunction

Identification of the CEC population from individual peripheral blood mononuclear cells (PBMCs) samples was carried out with high stringency using a panel of established CEC immunophenotypic markers (**Fig. 1a**). We first gated for the CD45^-^/CD31^+^ population to isolate CECs along with bone marrow-derived endothelial progenitor cells (EPCs) and platelets, while ruling out CD45^+^ leukocytes that also expressed the endothelial marker CD31 (PECAM1) (Duda, Cohen et al., 2007). This population was further subtyped based on the progenitor marker, CD133, to rule out EPCs. Finally, a nucleic acid stain was used to distinguish anucleate platelets from nucleated CECs defined here by a combined immunophenotypic profile of CD45^-^/CD31^+^/CD133^-^/DNA^+^ (Burger & Touyz, 2012, Duda et al., 2007, Yu, Lee et al., 2013). Identified CECs were further characterized for the expression of markers of endothelial cell activation, namely intercellular adhesion molecule 1 (ICAM1), P-selectin (SELP) and fractalkine (CX3CL1), that are integral for the processes of leukocyte adhesion, platelet aggregation and trans-endothelial migration respectively (Goncharov, Nadeev et al., 2017, Johnson & Jackson, 2013).

**Figure 1:**
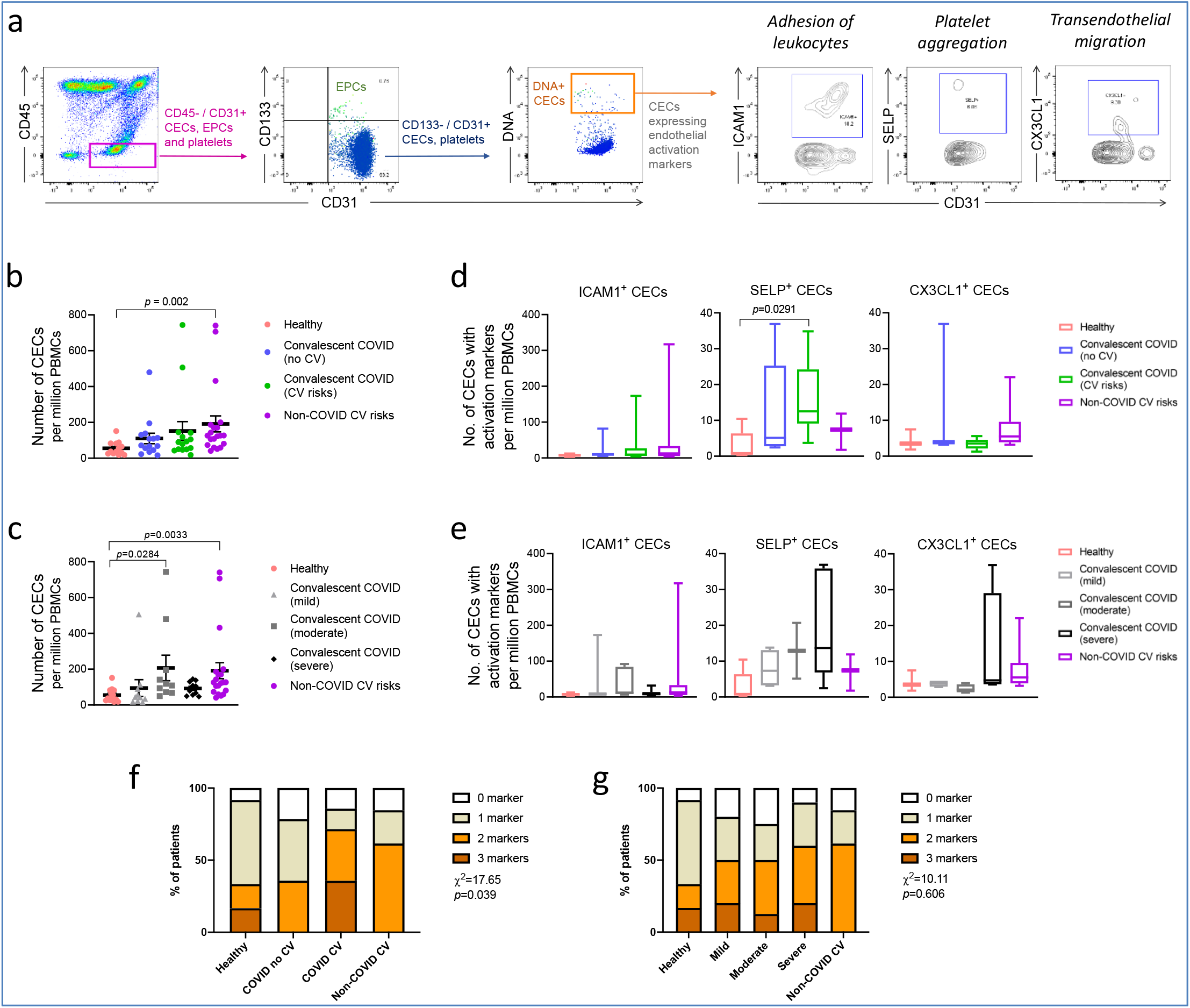
Enumeration and characterization of circulating endothelial cells from COVID-19 convalescents (n = 30), non-COVID-19 patients (n = 20) and healthy participants (n = 13). **(a)** Using flow cytometry, CEC populations were gated from PBMCs using a strategy involving positive (nuclear stain and CD31) and negative (CD45 and CD133) markers before characterization with three separate markers of endothelial activation. **(b) and (c)** Scatterplot visualization of the number of CECs per million PBMCs identified from each sample with mean and standard error of mean for each group shown. **(d) and (e)** Boxplots extending from 25^th^ to 75^th^ percentile with bar showing mean and whiskers indicating the minimum and maximum number of CECs per million PBMCs from each group staining positive for endothelial activation markers ICAM1, SELP or CX3CL1. Kruskal-Wallis test was performed in **(b-e)** to test for difference between the groups with Dunn’s multiple comparison test carried out for pairwise testing post-hoc. **(d)** The frequency of patients in each group with CECs staining positive for endothelial activation markers, ICAM1, SELP or CX3CL1 are shown here. **(f) and (g)** Cumulative analysis of patient frequency data of CECs staining positive for endothelial activation markers, ICAM1, SELP and/or CX3CL1. Chi-square goodness of fit test was performed to assess for difference in frequencies between groups in **(f)** and **(g)**.

CECs are typically rare in healthy individuals, occurring at between 0.0001% and 0.01% of the mononuclear cell population in peripheral blood (Khan, Solomon et al., 2005). Postulated as being shed from damaged vessels, elevated CEC numbers in the peripheral blood have been demonstrated as a cell-based biomarker for vascular dysfunction across a range of diseases including cardiovascular disease, preeclampsia, chronic kidney failure and sickle cell anemia (Goon, Boos et al., 2005). We enumerated and compared the mean CEC number per million PBMC between healthy individuals, convalescent COVID-19 patients, convalescent COVID-19 patients with cardiovascular risks and patients with cardiovascular risk but no history of COVID-19. While statistical significance was only found for the difference in mean CECs between healthy (*M* = 55.43, *SEM* = 10.64) and non-COVID-19 cardiovascular risk groups (*M* = 191.50, *SEM* = 44.77, *p* = .001), we observed a trend of more than two folds higher mean CEC numbers in both convalescent COVID-19 groups relative to healthy individuals (**Fig. 1b**). Between the two convalescent COVID-19 groups, patients with cardiovascular risk had higher mean CEC numbers (*M* = 153.75, *SEM* = 51.55) than patients without cardiovascular risk (*M* =116.21, *SEM* =28.17). These observations are consistent with our hypothesis that COVID-19 patients, especially those with pre-existing cardiovascular risk, may display persistent signs of vascular dysfunction even after recovery from COVID-19.

To ascertain the consequence of COVID-19 on endothelial dysfunction, we regrouped the convalescent COVID-19 patients by their disease severity during acute infection (**Fig. 1c**). We found that both convalescent COVID-19 patients who recovered from moderate symptoms (*p* = .0284) and non-COVID-19 patients with cardiovascular risks (*p* = .0033) had significantly higher numbers of CECs than healthy controls. It was surprising that convalescent COVID-19 patients from severe symptoms did not result in significantly higher CEC counts, probably due to limited representations in our current sample pool. Collectively, **Figures 1b and 1c** demonstrate the potential impact of COVID-19 and cardiovascular risk factors on endothelial injury, as indicated by the number of CECs in circulation. In **Figure 1d**, we further performed molecular profiling of CECs for endothelial activation markers. Our data revealed significant difference between the mean number of SELP^+^ CECs in convalescent COVID-19 patients with cardiovascular risk and healthy individuals (*p =* .0291). When we analyzed their endothelial activation hallmarks by disease severity (**Fig. 1e**), we observed a trend that convalescent COVID-19 patients who recovered from severe symptoms had the highest number of SELP+ CECs, though insignificant. As seen in the spread of data in **Figures 1d and 1e**, large inter-individual variations in CEC numbers was expected and could obscure observations of statistically significant differences in mean values. To overcome this, we compared the proportion of patients with regard to the numbers of activation markers expressed by CECs in each group and a chi-square goodness-of-fit test was carried out to assess if the percentages of patients were similar across the groups. We found a significant relationship between the number of endothelial activation markers expressed by CECs and the disease status of patients, *X*^2^ (9, N = 53) = 17.65, *p* = .03, where convalescent COVID-19 patients compounded with cardiovascular risk had the most pronounced endothelial activation hallmarks (**Fig. 1f**), more than those having either a history of COVID-19 alone or cardiovascular risk without COVID-19. Conversely, COVID-19 severity did not render significantly different extent of endothelial activation hallmarks (**Fig. 1g**). These findings demonstrate that COVID-19 may act in concert with comorbidities of hypertension and diabetes to intensify risks of future vascular complications due to elevated endothelial activation state.

### Persistent cytokine production in convalescent COVID-19 patients

Multiplex microbead-based immunoassay was performed to determine the cytokine levels in COVID-19 patient plasma during hospital admission and after discharge. To investigate the association between cytokine responses and cardiovascular risks, cytokine and chemokine levels were compared between COVID-19 patients with and without cardiovascular risk during the acute and convalescent phases of infection (**Fig. 2a**). We found that COVID-19 patients with cardiovascular risk had lower levels of growth factors BDNF (median concentration, 22.70 pg/mL *vs*. 38.57 pg/mL, *p* = .015), PDGF-BB (median concentration, 31.08 pg/mL *vs*. 62.51 pg/mL, *p* = .023) and PIGF-1 (median concentration, 30.69 pg/mL *vs*. 1.49 pg/mL, *p* = .040) compared to patients without cardiovascular risk at the early acute phase of infection (first plasma sample collected after hospital admission, at a median 10 days after symptom onset, IQR 7-15) (**Fig. 2b**). These growth factors are known to promote vascular function (Alomari, Khabour et al., 2015, Brown, Hong et al., 1995, Carmeliet, Moons et al., 2001) and our observations suggest that the endothelial dysfunction underlying patients with cardiovascular risk may impede vascular repair following injury during the acute phase of virus infection. Interestingly, the plasma levels of proinflammatory IL-1β (median concentration, 4.95 pg/mL *vs*. 3.51 pg/mL, *p* = .026), IL-17A (median concentration, 3.12 pg/mL *vs*. 0.09 pg/mL, *p* = .003), IL-2 (median concentration, 48.82 pg/mL *vs*. 27.07 pg/mL, *p* = .007) and RANTES (median concentration, 105.00 pg/mL *vs*. 72.47 pg/mL, *p* = .037) were significantly higher in patients with cardiovascular risk than those without (Fig. 2b) during the early convalescent phase (first plasma sample collected after hospital discharge, median 7 days post hospital discharge, IQR 3-12). Notably, the levels proinflammatory cytokines such as IL-1β, IL-17A, IL-2 and RANTES remained elevated in COVID-19 patients at the early convalescent phase, particularly in those with prior cardiovascular risk, compared to healthy controls (**Fig. 2b**). Result from simple linear regression analysis showed that age was significantly associated with the presence of cardiovascular risks in our COVID-19 patients. After adjustment for age by multiple linear regression analysis, levels of RANTES at the early convalescent phase was significantly associated with cardiovascular risks (**Supplementary Table 1**).

**Figure 2:**
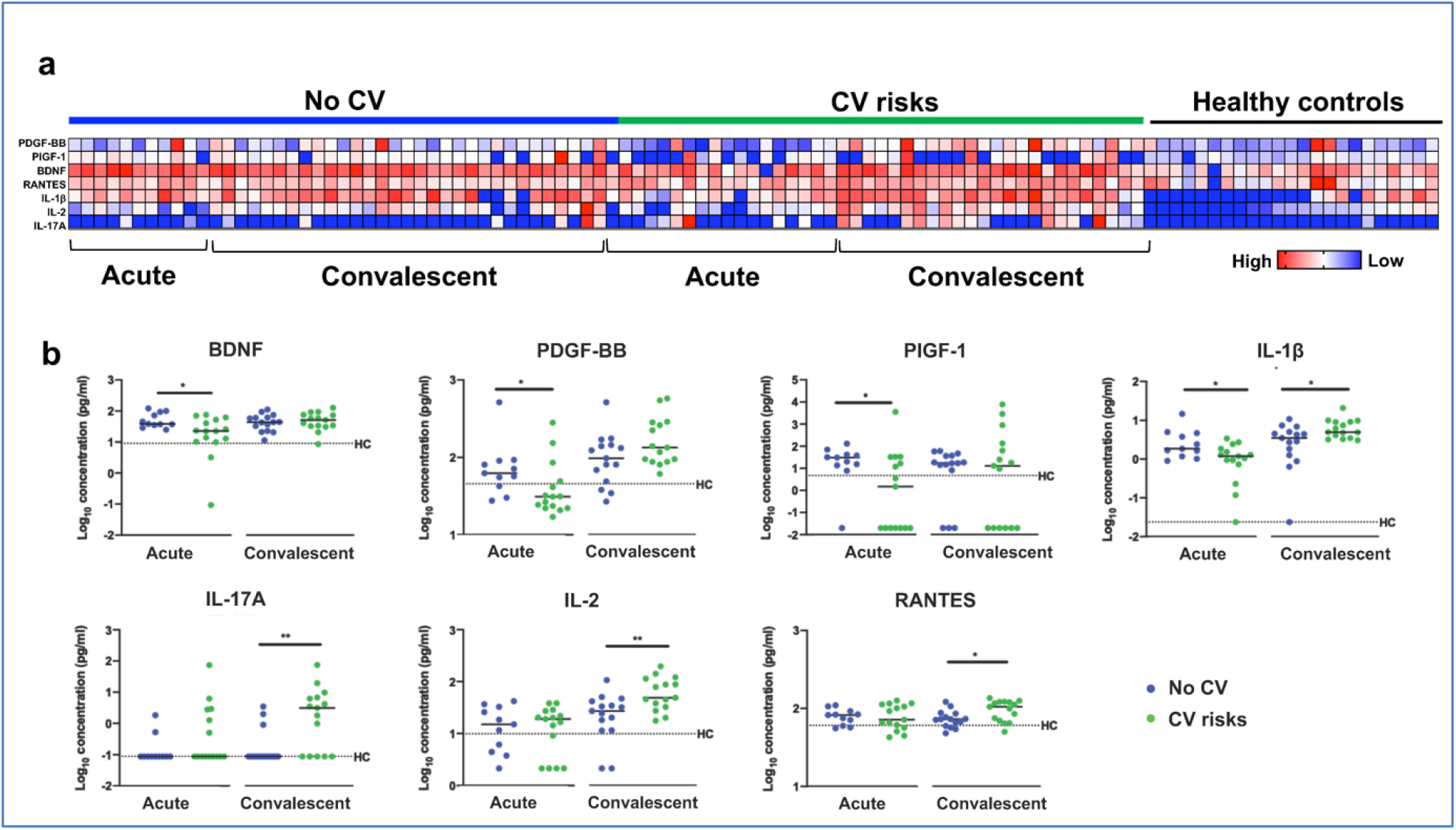
Plasma cytokine levels of COVID-19 patients with and without cardiovascular risk factors during the acute and convalescent phases of infection. Concentrations of 45 immune mediators were quantified using a 45-plex microbead-based immunoassay. **(a)** Heatmap of immune mediator levels in plasma samples of patients with (n = 15) and without (n = 15) cardiovascular risk factors at both acute (median 10 days post-illness onset), convalescent (0 to 90 days post-hospital discharge) phases of SARS-CoV-2 infection, and non-COVID-19 healthy controls. Each color represents the relative concentration of a particular analyte. Blue and red indicates low and high concentration, respectively. **(b)** Profiles of significant immune mediators of COVID-19 patients with and without cardiovascular risk factors at for first acute are illustrated as scatter plots. Cytokine levels in plasma fraction samples from first collection timepoint during hospital admission (acute, median 10 days post-illness onset) and discharge (convalescent, median 7 days post-hospital discharge) were compared. Mann-Whitney U were performed on the logarithmically transformed concentration (*p < 0.05; **p < 0.01). Immune mediator levels for healthy controls (n = 23) are indicated by the black dotted line. Patient samples with concentration out of measurement range are presented as the value of logarithm transformation of Limit of Quantification.

### Correlative studies implicating endothelial dysfunction with persistent cytokine production

To understand whether persistent immune activation may impact on endothelial dysfunction, we performed correlation analysis of cytokine levels with the aforementioned CEC attributes. Interestingly, CEC attributes from convalescent patients without prior cardiovascular risk correlated significantly with a greater number of cytokines than those with cardiovascular risk (**Fig. 3**). This may suggest that persistent cytokine production contributed primarily to endothelial injury (CEC counts) and activation hallmarks (CX3CL1+ and SELP+) in convalescent patients without previously known risk factors. On the other hand, more pronounced endothelial injury and activation in those with cardiovascular risk would have been attributed to their preexisting cardiovascular risk factors, on top of aftermath of COVID-19. Among the positively correlated cytokines in the cardiovascular risk group (**Fig. 3**), MIP-1α (CCL3) related to a chemoattractant for leukocytes, along with IL-17A,(Williams, Huang et al., 2019) IL-8(Apostolakis, Vogiatzi et al., 2009) and IL-18(Gerdes, Sukhova et al., 2002) known to evoke activation of endothelial cells during atherogenesis, may suggest chronic development of atherosclerotic plaques already in these individuals.

**Figure 3:**
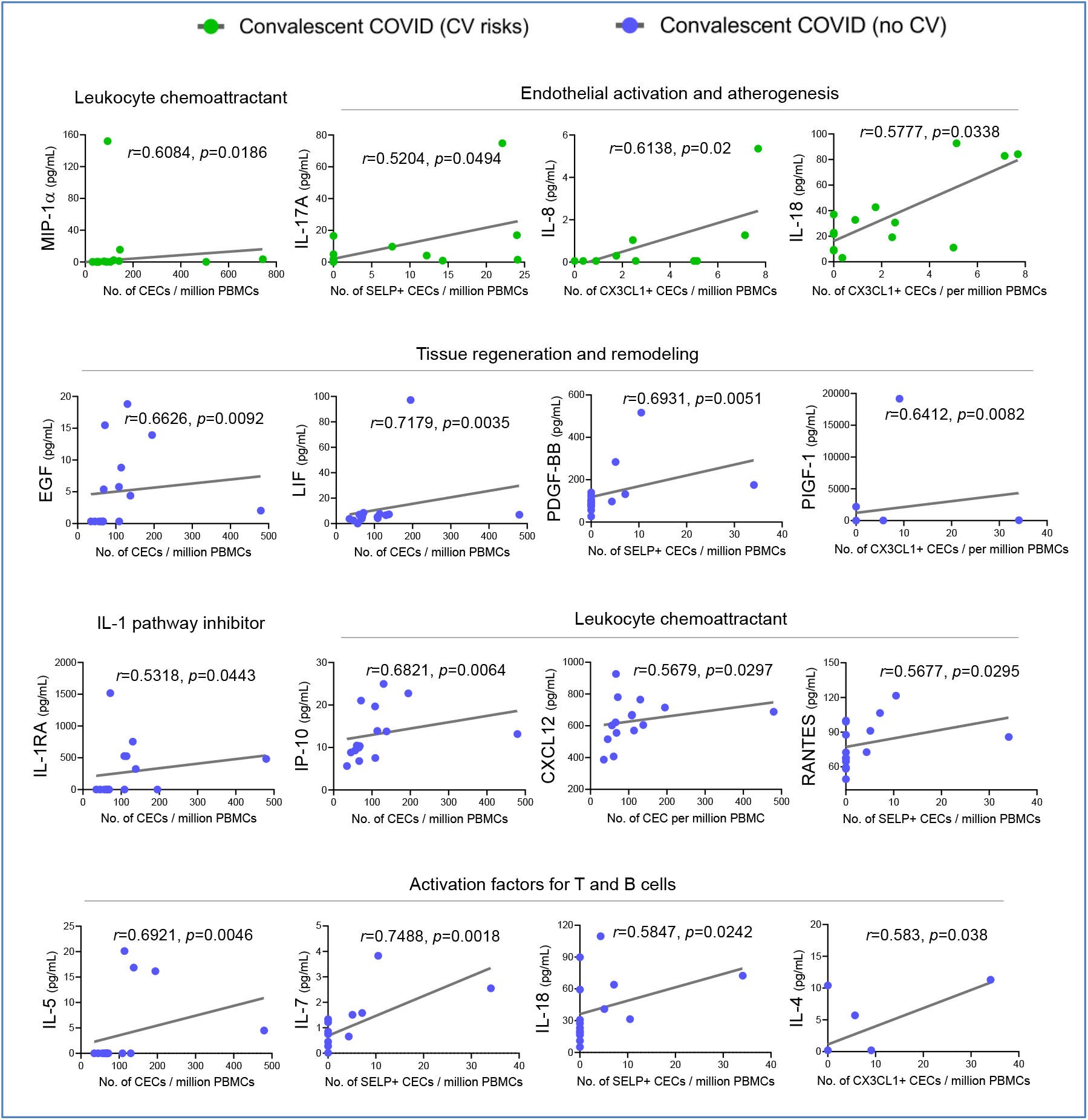
Significantly correlated cytokines with CEC attributes in COVID-19 convalescent patients. Spearman rank correlation coefficients were calculated to assess associations between the level of cytokines and CEC attributes in terms of mean numbers of CECs, SELP^+^ CECs or CX3CL1^+^ CECs. Spearman’s correlation coefficient r and *p* values (two-tailed test) were shown in plots.

For the convalescent patients without prior risk factors, the measures of CEC attributes reflected greater sensitivity to cytokine-driven endothelial dysfunction that could signify a more direct consequence of COVID-19. Further protein-protein interaction network analyses with STRING (Search Tool for the Retrieval of Interacting Genes/ Proteins) highlighted the direct and indirect interactions between the significant cytokines and their functional associations with biological processes involved in inflammatory response, vasculature development, angiogenesis, leukocyte chemotaxis, B cell proliferation and T cell activation (**Supplemental Fig**.**1**). Among the positively correlated cytokines (**Fig. 3**), we observed growth factors associated tissue regeneration and remodeling, namely EGF, LIF, PDGF-BB and PIGF-1 (PGF), indicating possibility of adaptive angiogenesis taking place as a response to preceding damages caused by viral infection and cytokine overdrive. IL-1RA, an IL-1 pathway inhibitor, could be an inflammation resolving mediator post infection. Conversely, IP-10 (CXCL10) is known to limit angiogenesis (Bodnar, Yates et al., 2006). This collective turnover of endothelial cells during blood vessel remodeling may result in elevated number of CECs. A number of significantly correlated cytokines are chemokines known to induce endothelial activation and promote chemotaxis of leukocytes to vascular endothelium. Chemotactic factors such as IP-10 (CXCL10), CXCL12 and RANTES (CCL5) regulate adhesion and transmigration of T lymphocytes, monocytes and/or neutrophils through endothelial barrier (Sokol & Luster, 2015). Our data resonated with the findings from a study on high-cardiovascular-risk patient cohort, where RANTES correlated positively with endothelial injury marker, and that increased perivascular expression of RANTES was associated with vascular accumulation of T cells (Mikolajczyk, Nosalski et al., 2016). Moreover, RANTES was specifically correlated with SELP+ CEC (*r* = 0.5677, *p* = 0.0295), indicating that activated platelet-derived RANTES may work in concert with endothelial SELP to mediate platelet aggregation and trigger coagulation cascade. In partial correlation analyses, a majority of these CEC-cytokines correlations were sustained even after accounting for age (**Supplemental Table 2**).

Overall, CEC attributes of convalescent COVID-19 patients correlated with a vast majority of differentiation and activation factors associated with T cells and B cells (IL-4, IL-5, IL-7, IL-17A, IL-18, MIP-1α and RANTES) (Turner, Nedjai et al., 2014) implicating a broad adaptive immune response with endothelial dysfunction. Besides being a target of the immune mediators, we recognize that endothelial cells, once activated, could also be a source of these cytokines, which in turn could activate their immune counterparts.

### Endothelial-immune crosstalk

Our ability to measure activation markers directly on CECs motivated us to better understand the receptor-receptor and chemokine-receptor interactions between activated endothelial cells with putative immune subpopulations in COVID-19. We performed data mining of published single-cell transcriptomic datasets on PBMCs from healthy participants, mild and severe COVID-19 patients (Wilk, Rustagi et al., 2020), and COVID-19 patients with or without cardiovascular disease (Schulte-Schrepping, Reusch et al., 2020). Then we re-analyzed the expressions of counter receptors (i.e. *ITGAL, SELPLG* and *CX3CR1*) to our endothelial activation markers (i.e. ICAM1, SELP and CX3CL1 respectively), in order to identify the potential immune interactors with activated endothelial cells. We found that those counter receptors were most pronouncedly expressed by CD8+ T cells, natural killer (NK) cells and to some extent monocytes (**Fig. 4a and Supplemental Figure 2**). In Wilk *et al*. dataset, the expression of *CX3CR1* was intensified in mild and severe COVID-19 patients than healthy participants (**Fig. 4a**). The relationship between counter receptors expression and COVID-19 disease was even more marked in Schulte-Schrepping *et al*. dataset with a higher proportion of NK or both CD8+ and NK cells expressing all three counter receptors found in mild and severe COVID-19 samples, respectively, regardless of comorbidity with cardiovascular disease (**Supplemental Figure 2)**. As those counter receptors generally marks cytotoxic effector lymphocytes in peripheral blood (Nishimura, Umehara et al., 2002), we correspondingly found that CD8+ T cells and NK cells were the major cytotoxic populations expressing perforin and granzymes (**Fig. 4b**), especially in COVID-19 patients (**Supplemental Figure 2)**. Published immunophenotyping studies on convalescent COVID-19 patients are still limited. Several immunophenotyping studies have reported immune exhaustion phenotype and reduced antiviral cytokine production capability that were inversely correlated with COVID-19 severity during acute infection (summarized in **Supplemental Table 3**). The reason underlying such lymphopenia remains unclear and could be either due to an impaired maturation possibly due an increased level of inflammatory cytokine such as IL-6 (Cifaldi, Prencipe et al., 2015, Mazzoni, Salvati et al., 2020) or the recruitment of the mature T and NK cells into the lungs or other organs (Liao, Liu et al., 2020, Varga et al., 2020, Xu et al., 2020). Conversely, in many convalescent patients and even in those who had recovered from mild symptoms, broad CD8+ T-cell response against multiple SARS-CoV-2 proteins mediated by functional effector and memory T cell populations were detected, suggesting a key role for T cells in protective immunity in COVID-19 convalescents (**Supplemental Table 3**). Intriguingly, a recent immunophenotyping study using high-dimensional flow cytometry reported a higher proportion of activated CD8+ T cells in healthy participants and to some extend recovered individuals than in COVID-19 patients (Mathew, Giles et al., 2020). Therefore, we postulate that persistent cytokine production not only activates endothelial cells in convalescent COVID-19 patient but may promote direct interaction between activated endothelial cells and cytotoxic effector cells. Therefore, these individuals could be susceptible to further cytotoxicity-induced endothelial injury.

**Figure 4:**
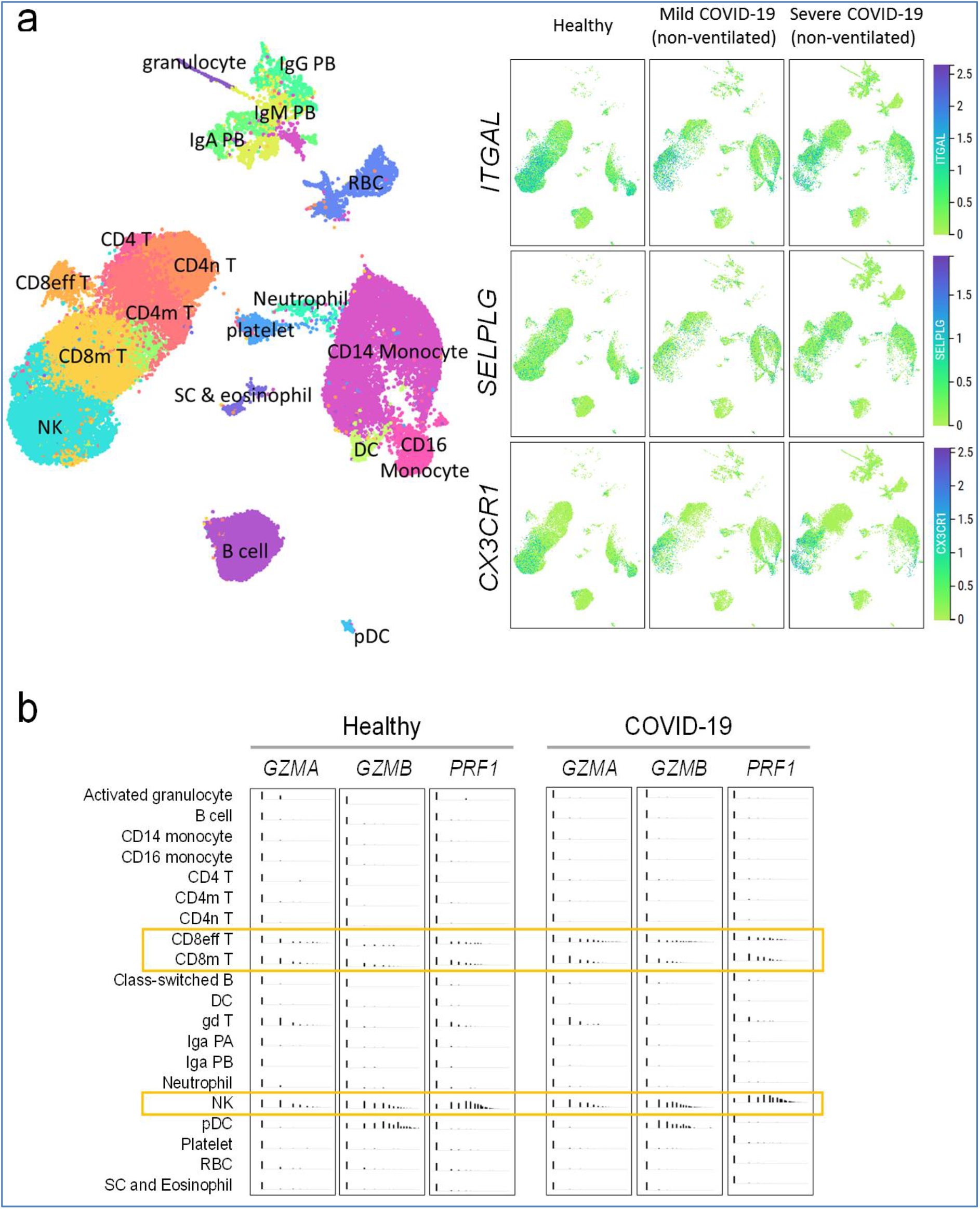
Immune interactors of activated endothelial cells. **(a)** UMAP representations of immune cell populations from healthy participants and COVID-19 patients annotated by cell types (left) and differential expressions of counter receptors ITGAL, SELPLG and CX3CR1 which are known to interact with surface molecules of activated endothelial cells (right). **(b)** Distribution of the expressions of cytotoxic genes GZMA, GZMB and PRF1 across immune cell populations.

## DISCUSSIONS

The rapid rise of COVID-19 “long-haulers” with lingering symptoms post-infection, or recovered individuals who suffer from sudden cardiovascular events, are expected to lead to a worldwide shift in disease burden in the near future that would continue to place tremendous strain on our healthcare systems. The understanding of COVID-19 intermediate and long-term impact on vascular health is still lacking. Thus, we performed vascular phenotyping through the analysis of CECs in convalescent COVID-19 patients. Our findings reveal that endothelial dysfunction was most pronounced in convalescent COVID-19 patients with pre-existing cardiovascular risk factors (i.e. hypertension, hyperlipidemia, diabetes). Furthermore, markers of endothelial injury and activation in convalescent COVID-19 patients were significantly correlated with a number of proinflammatory cytokines that persisted into convalescent phase, suggesting that recovered individuals may be vulnerable to vascular instability.

About 2.5% of convalescent COVID-19 patients are found with thrombosis (including arterial and venous events), 30 days post discharge (Patell, Bogue et al., 2020). As endothelial dysfunction often precedes serious thrombotic complications and ischemic damages to organs, we detected subclinical endothelial changes using CECs and associated activation markers. Most previous studies focused on enumeration of vascular progenitor and endothelial cells in the blood. Here, even with a stringent marker profile of CD45^-^/ CD133^-^/ DNA^+^/ CD31^+^ to identify live CECs, it is common that CEC numbers present high inter-individual variability. Circulatory release of CECs may be altered if some patients had been treated with metformin or drugs that may confer some vasculoprotective effects,(Ahmed, Rider et al., 2016) potentially leading to fewer endothelial cells being shed off the vascular lining. Beyond enumeration of CECs, we further measured ICAM1, SELP and CX3CL1 on CECs, expressions of which would suggest proinflammatory and procoagulant state of the endothelial cells originating from sites of vascular injury. In comparing with non-COVID-19 patients with cardiovascular risk factors (i.e. hypertension, diabetes, coronary artery disease), the compounded effects of COVID-19 with prior cardiovascular risk factors rendered convalescent patients with more SELP-expressing CECs, and the greatest proportion within this group of individuals possessed all three endothelial activation markers. This suggests a potential role of endothelial damage in the pathogenesis of post infection thrombosis.

The sequence of events leading to post COVID-19 complication has not been fully elucidated, but likely a vicious cycle caused by vascular leakage, activation of coagulation pathway and inflammation (Teuwen, Geldhof et al., 2020). Breach of endothelial barriers might have occurred early due to impairment of ACE2 upon SARS-CoV-2 entry, indirectly perturbing the kallikrein–bradykinin pathway,(Garvin, Alvarez et al., 2020) thus resulting in vascular hyperpermeability. Direct SARS-CoV-2 infection impair the renin– angiotensin pathway (Ingraham, Barakat et al., 2020). Angiotensin II is known to promote vascular inflammation and stimulates endothelial secretion of CXCL10 which in turn acts in an autocrine manner to promote angiotensin II-induced endothelial cell apoptosis (Ide, Hirase et al., 2008). Here, our cytokine profiling confirmed that cytokine production remained heightened post-infection. Of interest, the CEC attributes of convalescent patients correlated significantly with a number of proinflammatory cytokines, indicating that prolonged overactive state of the immune system was implicated with endothelial dysfunction, especially for those without prior cardiovascular risk factors. Hence, viral-induced damage exposes tissue factors on endothelial cells which are activated by persistent cytokines to produce chemoattractants in recruiting leukocytes to endothelial cells. Together with activated monocytes and platelets, endothelial cells mediate extrinsic coagulation pathway, leading to blood clotting.

In our study, a number of chemokines and activated T cell-associated cytokines (IL-1β, IL-2, IL-17A, and RANTES) remained elevated post-infection, reflecting a broad adaptive immune response. We noted much heterogeneity in the immune profiles of COVID-19 patients where one study described no substantial expression of proinflammatory cytokine genes by T cells, NK cells or monocytes, to another that found higher proportion of cytotoxic CD8+ T cells in severe COVID-19 individuals (summarized in **Supplemental Table 3**). Similarly, we also noted differences in the expression of counter receptor and cytotoxicity associated genes between the two scRNA datasets that we analyzed although both studies supported higher proportion of CD8+ and NK cells expressing counter receptor and cytotoxicity associated genes in COVID-19 samples. Importantly, no difference in the expression of counter receptor and cytotoxicity associated genes were noted in COVID-19 patients with or without cardiovascular risk. While the nature of the cardiovascular risk in Schulte-Schrepping *et al* dataset is not elaborated, it supports that factors other than immune cytotoxicity may impact vascular health. Correspondingly, SARS-CoV-2 reactive T cells were described in COVID-19 convalescents and even unexposed individuals (Grifoni, Weiskopf et al., 2020). We postulate that activated endothelial cells express receptors and ligands which regulate interactions with the counter receptors on functional effector and memory T cell populations, making activated endothelial cells a potential target of cytotoxic effector lymphocytes. Immune cell-mediated endothelial injury has been observed in other viral infections, including Ebola, human cytomegalovirus infection and malaria, where T cells recruited to infected site by inflammation could induce apoptosis of infected endothelial cells, causing vascular leakage (Claser, Nguee et al., 2019, van de Berg, Yong et al., 2012, Wolf, Kann et al., 2015, Yang, Duckers et al., 2000). Likewise, in non-infectious diseases such as acute coronary syndrome and systemic sclerosis, cytotoxic CD4+ T cells were capable of causing damages to endothelial cells (Maehara, Kaneko et al., 2020, Nakajima, Schulte et al., 2002). It is plausible that infected endothelial cells could be susceptible to direct cytotoxic T cell-mediated killing due to the immune overdrive in COVID-19. Subsequently, how persistent cytokine response and functional effector T cell populations could translate into endothelial dysfunction during convalescence warrant further investigation.

We are mindful of the limitations with this current study. First, we noted the major determinants of cardiovascular risks in convalescent COVID-19 and non-COVID-19 patients, but there might have been unmeasured confounding conditions that could contribute to some heterogeneity in our CEC and cytokine profiling. Second, convalescent blood samples were collected at various time points due to logistical constraints in recalling recovered patients on the same day post-infection. Finally, to understand post-COVID-19 complications, long-term phenotyping of vascular functions and immune profiles of convalescent patients is still lacking due to short term horizons from when these individuals were first infected. There is a critical need to monitor prospective cohorts of recovered individuals in order to establish the full spectrum of clinical courses of complications. Global efforts in this aspect are evident and are underway.

Hematologic assessment (i.e. thrombin generation, platelet activation studies, von Willebrand factor) is commonly performed to help manage convalescent patients who have been re-admitted for acute thrombosis. It would be valuable to risk stratify convalescent patients before adverse events happen. Our profiling of CECs may serve as a form of vascular surveillance to complement hematologic assessment in identifying early molecular and endothelial changes in high-risk individuals. Analysis of CECs may detect subclinical changes in the vasculatures and be able to provide accurate and easily transferable endpoints in clinical assessment.

In summary, managing the aftermath of COVID-19 is an imperative. Endothelial instability may be a key mechanism underpinning the development of post-infection vascular complications. Clinical trials in preventive therapy for vascular complications may be needed.

## MATERIALS AND METHODS

### Study design, participants and clinical data collection of convalescent COVID-19 patients

Convalescent COVID-19 individuals were recalled from the PROTECT study which is a prospective observational cohort study at three public hospitals in Singapore (the National Centre for Infectious Diseases, National University Hospital and Changi General Hospital). Written informed consent was obtained from participants who provided clinical data and biological samples. Study protocols were approved by ethics committees of the National Healthcare Group (2012/00917).

### Biological sample collection and processing for convalescent COVID-19 patients

The electronic medical records of patients enrolled in the PROTECT study were reviewed and their data entered onto a standardized collection form adapted from the International Severe Acute Respiratory and Emerging Infection Consortium’s case record form for emerging severe acute respiratory infections. Serial blood samples were collected during hospitalization and post-discharge. Blood samples were processed as previously reported (Young, Ong et al., 2020b, Young et al., 2020c). Ficoll centrifugation was performed to retrieve PBMC fraction for downstream CEC characterization. Plasma samples were separately stored for subsequent cytokine profiling.

### Study design, participants and clinical data collection of healthy participants and non-COVID-19 patients with cardiovascular risk factors

Healthy participants and non-COVID-19 patients with cardiovascular risk factors were obtained from the Cardiac Ageing study (Koh, Gao et al., 2018) which is a prospective observational cohort study performed at the National Heart Centre Singapore. The current study sample consisted of healthy participants who had no known cardiovascular disease or cerebrovascular disease or cancer. We included non-COVID-19 patients with cardiovascular risk factors for this comparison. All participants were examined and interviewed on one study visit by trained study coordinators. Participants completed a standardized questionnaire that included medical history and coronary risk factors. Sinus rhythm status was ascertained by resting electrocardiogram. Clinical data were obtained on the same day as biological sample collection.

### Biological sample collection and processing for healthy participants and non-COVID-19 patients with cardiovascular risk factors

Antecubital venous blood samples were taken from participants on the same day. After collection, the blood samples were immediately placed on ice for transportation and were processed within six hours to obtain buffy coat samples, which were subsequently cryopreserved.

### Vascular phenotyping by flow cytometric analysis

Study protocols were approved by ethics committees of the Nanyang Technological University Singapore (IRB-2020-09-011). PBMC samples were washed with DPBS (Dulbecco’s Phosphate-Buffered Saline, Hyclone, SH30028.02) + 1% BSA (Bovine Serum Albumin, Hyclone, SH30574.02), and then resuspended in 100 μl of DPBS + 1% BSA for antibody staining (**Supplemental Methods & Materials**). Staining was carried out in the dark for 10 minutes at room temperature, followed by 20 minutes at 4°C on an analog tube rotator. After staining, cells were rinsed and resuspended in DPBS + 1% BSA for downstream flow cytometry analysis. Flow cytometry was performed using BD LSRFortessa X-20 (BD Biosciences) and data acquisition was performed on FACSDiva software, version 8.0.1 (BD Biosciences). Spectral overlap between INDO-1, APC, PE, PE-Cy7, AF488 and BV711 channels was calculated automatically by the FACSDiva software after measuring single-color compensation controls from pooled PBMCs. Optimal compensation was achieved using compensation control beads (Anti-Mouse Ig, κ/Negative Control Compensation Particles Set, 552843) together with corresponding conjugated antibodies. Acquired data were analyzed using FlowJo software, version 10.7.1. Analysis of each patient typically included between 50,000 – 200,000 PBMCs depending on sample availabilities. CECs were detected by a combined immunophenotypic profile of CD45-/CD31+/CD133-/DNA+ and were further characterized for the expressions of ICAM1, SELP and CX3CL1. Attributes such as CECs or ‘activated’ CECs were represented as cells per million of PBMCs in Figure 1.

### Data mining of published immune single-cell transcriptomes of COVID-19

Single cell transcriptomic dataset published by Wilk et al., 2020 and Schulte-Schrepping et al. were re-analyzed using the cellxgene platform (https://cellxgene.cziscience.com/d/Single_cell_atlas_of_peripheral_immune_response_to_SARS_CoV_2_infection-25.cxg/). Briefly, the uniform manifold approximation and projection (UMAP) visualization was annotated using the metadata included using the “cell_type_fine” taxonomy. The expression data was obtained by interrogating the individual expression of the genes in the UMAP (*CX3CR1, ITGAL, SELPLG)* or across the various immune population included in the “cell_type_fine” taxonomy (*PRF1, GZMA and GZMB)*.

### Cytokine analysis by multiplex microbead-based immunoassay

Plasma samples were treated with 1% Triton™ X-100 solvent-detergent mix for virus inactivation (Darnell & Taylor, 2006) Cytokine levels in COVID-19 patient plasma across different acute and convalescent timepoints were measured with the Luminex assay using the Cytokine/Chemokine/Growth Factor 45-plex Human ProcartaPlex Panel 1 (ThermoFisher Scientific; **Supplemental Methods and Materials**). Patient samples with a concentration out of measurement range were assigned the value of the logarithmic transformation of the limit of quantification. Data analysis was done with Bio-Plex Manager 6.1.1 software.

### Statistics

Due to inter-individual heterogeneities of flow cytometric and cytokine data, nonparametric tests of association were preferentially used throughout this study unless otherwise stated. For flow cytometric data on CEC attributes, statistical differences between groups were calculated using Kruskal-Wallis test with Dunn’s multiple comparison post-test. Significant *p* values (< 0.05) were indicated on the graphs directly. To assess for differences in the frequencies of patients with CECs expressing endothelial activation markers, ICAM1, SELP or CX3CL1 and the cumulative frequencies for all markers, the number of patients per group were summarised in contingency tables. These were then analysed with Chi-square goodness of fit test with expected values generated from the data using GraphPad Prism, version 8.3.1.

For the cytokine analysis, Mann Whitney U tests were used to discern the differences in cytokine levels between the patients with or without cardiovascular risk factors. A multiple linear regression analysis was conducted to examine the association between plasma cytokines and the presence of cardiovascular risks in COVID-19 patients after adjustment for age. Heatmap and scatter plots were generated using GraphPad Prism version 8. Concentrations of immune mediators were scaled between 0 and 1 for visualization in the heatmap. Protein-protein interaction networks of the CEC associated cytokines were predicted and illustrated with Search Tool for the Retrieval of Interacting Genes/Proteins database (STRING) (version 11.0; available at: https://string-db.org). All the interactions between cytokines were derived from high-throughput lab experiments and previous knowledge in curated databases at a confidence threshold of 0.4. For correlative study between CEC attributes and cytokine levels, we selected non-parametric Spearman correlation with no assumption regarding value distribution. Non-linear regression and Spearman rank correlation coefficients were calculated to access associations between the level of serum cytokines and CEC attributes for convalescent COVID-19 patients with and without cardiovascular risk factors. The level of statistical significance was set at two-tailed test with *p* values of <0.05 to detect any significant associations between specific cytokines and CEC attributes. Statistical analyses for this correlative study were performed using GraphPad Prism software, version 8.3.1. To address potential age-related contributions to the correlations observed between CEC and plasma cytokines in Figure 3, non-parametric partial correlation coefficients based on Spearman’s rank correlations were determined for the identified correlated variables while controlling for age. The R package ‘ppcor’ was used to compute the coefficients, *p* values and test statistics in **Supplemental Table 2**. (Kim, 2015).

## Data Availability

Data are available as they are reported in the manuscript.

## STUDY APPOVAL

This study was approved by the Local Ethics Committee of the National Healthcare Group (2012/00917) and Nanyang Technological University Singapore Institutional Review Board (IRB-2020-09-011).

## ACKNOWLEDGEMENTS

We thank all clinical and nursing staff who provided care for the patients; staff in the Singapore Infectious Disease Clinical Research Network and Infectious Disease Research and Training Office of the National Centre for Infectious Diseases for coordinating patient recruitment. We also wish to thank Ms Siti Naqiah Amrun, Ms Rhonda Sin-Ling Chee, Mr Nicholas Kim-Wah Yeo, Mr Anthony Torres-Ruesta, Drs Chek Meng Poh, Cheryl Yi-Pin Lee, Guillaume Carissimo, Matthew Tay and Zi-Wei Chang from the Singapore Immunology Network (SIgN), for their help in isolating PBMCs and plasma fractions from the blood of COVID-19 patients. We are also grateful to Dr Danielle Anderson and her team at Duke-NUS, for their technical assistance in virus inactivation procedures with Triton-X-100™; Dr. Olaf Rötzschke, Dr. Bernett Lee, Wilson How and Norman Leo Fernandez from the Singapore Immunology Network (SIgN) Multiplex Analysis of Proteins (MAP) platform, for their assistance in running multiplex microbead-based immunoassay. We also thank Ms Manisha Cooray, Dr Chun-Yi Ng and Ms Natalie Yeo for their technical help during experimental optimization.

## FUNDING

This study was funded by the National Medical Research Council COVID-19 Research Fund (COVID19RF-001 and COVID19RF-060) and COVID-19 (project number H20/04/g1/006) research grants provided to Singapore Immunology Network (SIgN) by the Biomedical Research Council (BMRC), A*STAR. The SIgN flow cytometry, Multiple analyte platforms and Immunomonitoring Service Platform (ISP) were supported a grant (#NRF2017_SISFP09) from the National Research Foundation (NRF), Singapore. The team from Nanyang Technological University Singapore was funded by the Nanyang Assistant Professorship, Academic Research Fund grant (MOE2018-T2-1-042) from the Ministry of Education, Singapore, and an Industrial Alignment Fund Pre-Positioning grant (H18/01/a0/017) from Agency for Science, Technology and Research, Singapore. The team from National Heart Centre Singapore was supported by the National Medical Research Council of Singapore (NMRC/TA/0031/2015, MOH-000153, NMRC/OFIRG/0018/2016, NMRC/BnB/0017/2015, MOH-000358).

## AUTHOR CONTRIBUTIONS

NCID initiated the PROTECT cohort, NCID, NHCS, NUH and CGH collected the clinical samples and data, while NTU performed CEC phenotyping and data interpretation, and A*STAR’s SIgN performed the immunoassays. CC, DCL, LFPN and LR conceived the initial concept and designed the study. BEY and ASK collected the biological samples and clinical data. FWJC, SWF and YHC performed experiments and analyzed data. KXW and AS performed statistics and data mining. CC, SWF, KXW, AS wrote the manuscript and all authors approved the final manuscript. The order of co-first authors are assigned based on their experimental involvement, intellectual input and significance of findings.

## CONFLICT OF INTEREST

The authors declare that they have no conflict of interest.

## Notes

### Competing Interest Statement

The authors have declared no competing interest.

