## Supplemental information for "Convalescent COVID-19 patients are susceptible to endothelial dysfunction due to persistent immune activation"

### Supplemental Methods and Materials

***Antibody panel for detection of circulating endothelial cells***

| **Antibodies** | **Manufacturer (Cat. number)** |
|---|---|
| Hoechst 33342 Ready Flow™ Reagent | Invitrogen (R37165) |
| APC-conjugated monoclonal antibody against human CD133 | Biolegend (372806) |
| PE-conjugated monoclonal antibody against human CD45 | Biolegend (304008) |
| PE-Cy7-conjugated monoclonal antibody against human CD31 | Biolegend (303118) |
| AF488-conjugated monoclonal antibody against human CX3CL1/Fractalkine Chemokine Domain | R&D Systems (IC365G-100UG) |
| BV711-conjugated monoclonal antibody against human C54 | BD Biosciences (564078) |
| AF488-conjugated monoclonal antibody against human CD62P | Biolegend (304916) |

***Cytokine analysis by multiplex microbead-based immunoassay***

Plasma samples were treated with 1% Triton™ X-100 solvent-detergent mix for virus inactivation.[1] Cytokine levels in COVID-19 patient plasma across different acute and convalescent timepoints were measured with the Luminex assay using the Cytokine/Chemokine/Growth Factor 45-plex Human ProcartaPlex Panel 1 (ThermoFisher Scientific). The Cytokine/Chemokine/Growth Factor 45-plex Human ProcartaPlex™ Panel 1 panel included granulocyte-macrophage colony-stimulating factor (GM-CSF), epidermal growth factor (EGF), brain-derived neurotropic factor, beta-nerve growth factor (bNGF), basic fibroblast growth factor (FGF-2), hepatocyte growth factor (HGF), monocyte chemoattractant protein (MCP) 1, macrophage inflammatory protein (MIP) 1$\alpha$, MIP-1$\beta$, RANTES (regulated on activation, normal T cell expressed and secreted), chemokine (C-X-C motif) ligand (CXCL) 1 (GRO-$\alpha$), stromal cell-derived factor 1 (SDF-1$\alpha$), interferon (IFN) gamma-induced protein 10 (IP-10), eotaxin, IFN-$\alpha$, IFN-$\gamma$, interleukin (IL) IL-1$\alpha$, IL-1$\beta$, IL-1RA, IL-2, IL-4, IL-5, IL-6, IL-7, IL-8, IL-9, IL-10, IL-12p70, IL-13, IL-15, IL-17A, IL-18, IL-21, IL-22, IL-23, IL-27, IL-31, leukemia inhibitory factor (LIF), stem cell factor (SCF), tumor necrosis factor (TNF-$\alpha$), TNF-$\beta$, vascular endothelial growth factors A and D (VEGF-A, VEGF-D), platelet derived growth factor (PDGF-BB), and placental growth factor (PLGF-1). Standards and plasma from COVID-19 patients and healthy controls were incubated with fluorescent-coded magnetic beads pre-coated with respective antibodies in a black 96-well clear-bottom plate overnight at 4˚C. After incubation, plates were washed 5 times with wash buffer (PBS with 1% BSA (Capricorn Scientific) and 0.01% Tween (Promega). Sample-antibody-bead complexes were incubated with Biotinylated detection antibodies for 1 hour and washed 5 times with wash buffer. Subsequently, Streptavidin-PE was added and incubated for another 30 mins. Plates were washed 5 times again before sample-antibody-bead complexes were re-suspended in sheath fluid for acquisition on the FLEXMAP® 3D (Luminex) using xPONENT® 4.0 (Luminex) software. Internal control samples were included in each Luminex assays to remove any potential plate effects. Readouts of these samples were then used to normalize the assayed plates. A correction factor was obtained from the differences observed across the multiple assays and this correction factor was then used to normalize all the samples. Standard curves were generated with a 5-PL (5-parameter logistic) algorithm, reporting values for mean florescence intensity (MFI) and concentration data. The concentrations were logarithmically transformed to ensure normality. Patient samples with a concentration out of measurement range were assigned the value of the logarithmic transformation of the limit of quantification. Data analysis was done with Bio-Plex Manager 6.1.1 software.

### Supplemental Figure 1

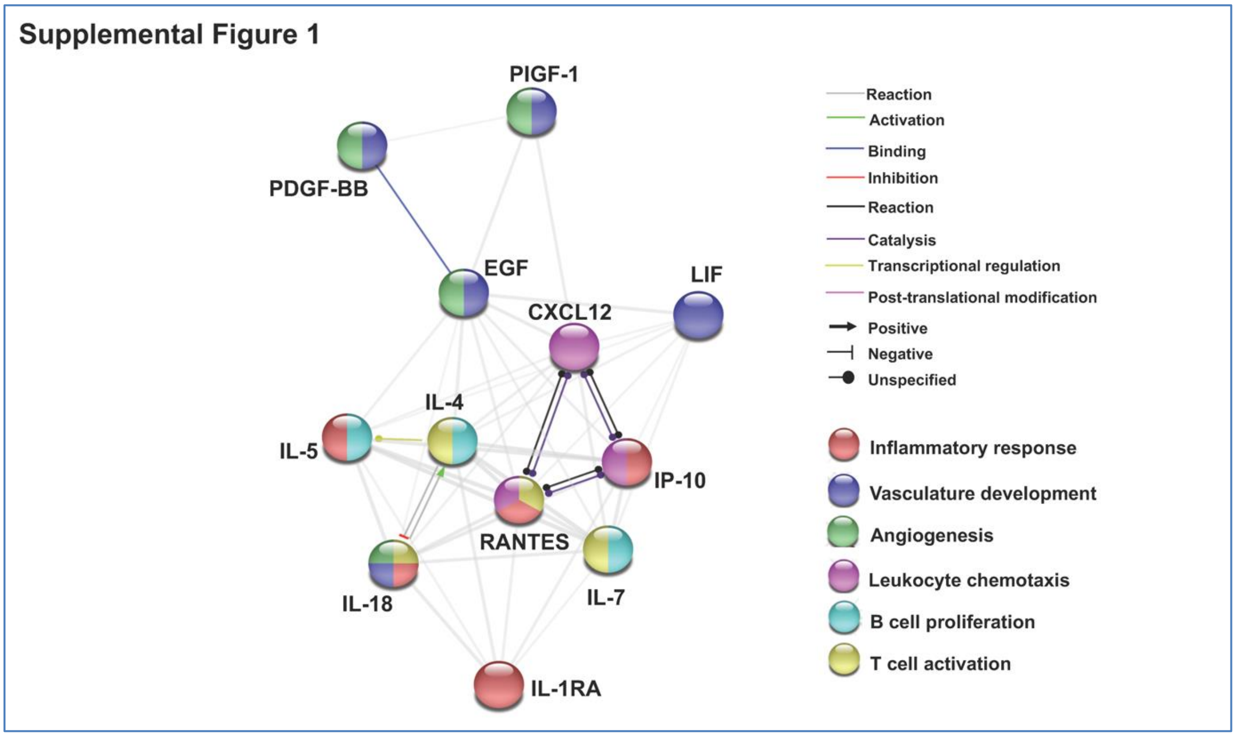


**Supplemental Figure 1: Network analysis of CEC associated cytokines in the convalescent COVID-19 patients without prior cardiovascular risk factors**. Interactive relationships between the cytokines or chemokines were determined by STRING (Search Tool for the Retrieval of Interacting Genes/ Proteins) analysis, with a confidence threshold of 0.4.

### Supplemental Figure 2

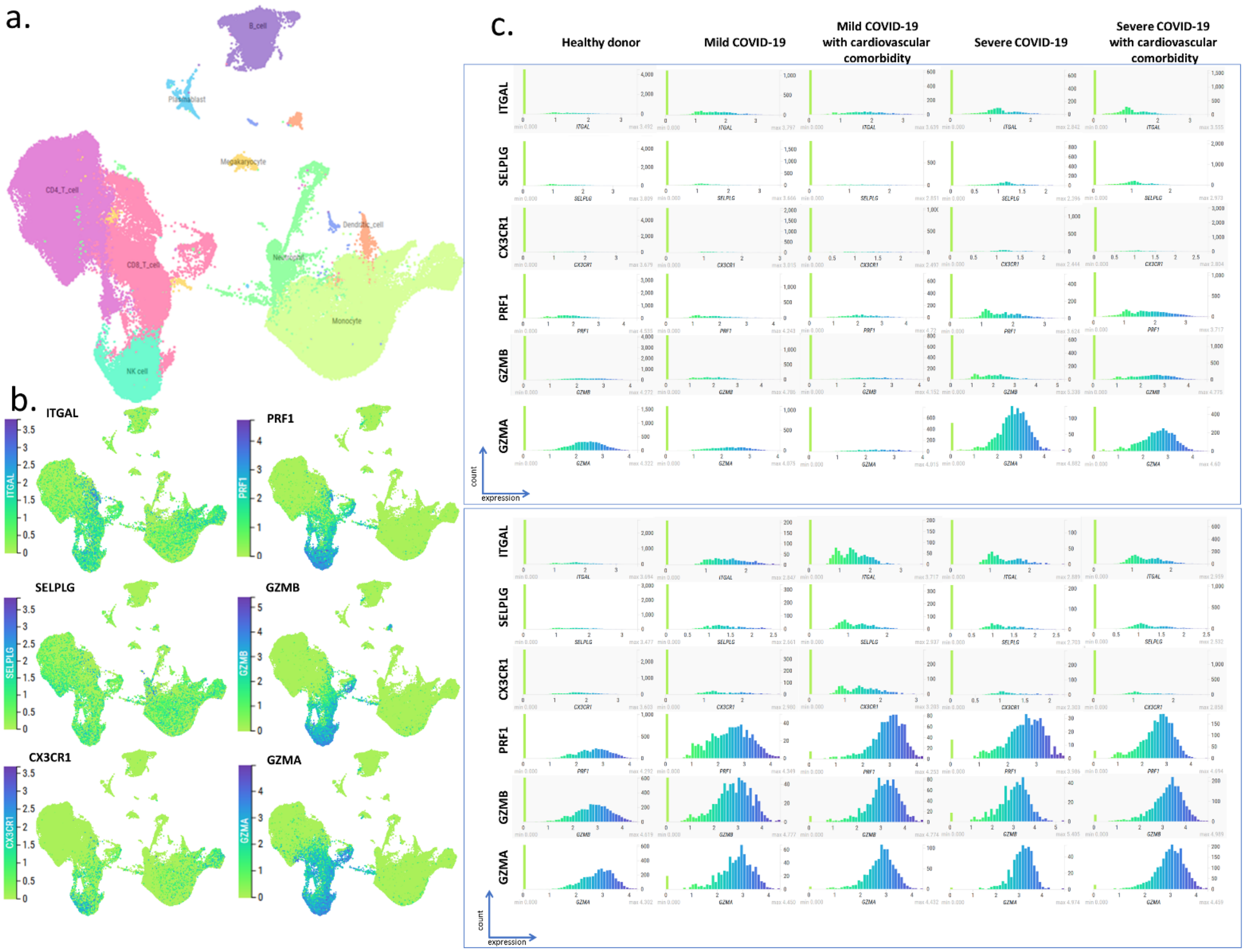


**Supplemental Figure 2: COVID-19 samples with or without cardiovascular comorbidity have a higher proportion of immune cells expressing counter receptor and cytotoxicity associated genes.**
**a)** Single cell transcriptomic dataset published by Schulte-Schrepping et al., 2020 were re-analyzed using the cellxgene platform hosted by Fastgenomics database (https://beta.fastgenomics.org/datasets/detail-dataset-952687f71ef34322a850553c4a24e82e#Cellxgene). The samples were annotated as mild (WHO 2-4) or severe (5-7) COVID-19 disease according to the WHO clinical ordinal scale and with or without cardiovascular commodity following publication Table S1 annotations. The UMAP representing immune cell populations compiled from healthy donor (n=21), mild with (n=4) or without (n=4) cardiovascular comorbidity and severe with (n=8) or without (n=2) cardiovascular comorbidity was annotated using the metadata included in the "cluster_labels_res.0.4els_res.0.4" taxonomy. For better clarity, the clusters of the same cell type were pooled together.
**b)** Expression of counter receptors (*ITGAL*, *SELPLG* and *CX3CR1)* and cytotoxicity associated genes (*PFR1, GMZB* and *GMZA*) across immune cell population. Both set of genes are mainly expressed by CD8+ and NK cells and to some extent monocyte.
**c)** Distribution of the expression of counter receptors and cytotoxicity associated genes in CD8+ cell (top) and NK (bottom) in mild and severe COVID-19 samples with or without cardiovascular comorbidity. Compared to healthy donors, mild and severe COVID-19 samples have a higher proportion of NK or both CD8+ and NK cells expressing counter receptors and cytotoxicity associated genes, respectively. No difference can be observed in presence or absence of cardiovascular comorbidity.

### Supplemental Table 1

Plasma cytokines and association with cardiovascular risk factors.

| | β (95% CI) | *p* value |
|---|---|---|
| Age | 12.33 (3.83-20.83) | **0.006** |
| **Early acute phase** | | |
| BDNF | *–18.16 (–45.07-8.75) | 0.176 |
| PDGF-BB | *–30.98 (–133.80-71.80) | 0.539 |
| PIGF-1 | *18.56 (–664.20-701.30) | 0.956 |
| IL-1β | *–1.56 (–4.14-1.02) | 0.225 |
| **Early convalescent phase** | | |
| IL-1β | *2.39 (–0.92-5.71) | 0.150 |
| IL-17A | *5.11 (–6.39-16.61) | 0.370 |
| IL-2 | *32.92 (–1.88-67.72) | 0.062 |
| RANTES | *21.18 (0.46-41.90) | **0.045** |

Bold indicates statistical significance at alpha level 0.05. [a] Linear regression models with plasma cytokines were adjusted for age.

### Supplemental Table 2

Partial correlation of plasma cytokines and number of CECs after controlling for age.

| Correlated variables from Figure 3 | | Partial correlation coefficient (*r*) | *p* value* | Test statistic |
|---|---|---|---|---|
| ***Convalescent COVID with CV risks*** | | | | |
| MIP-1α | CEC | 0.595 | **0.032** | 2.457 |
| IL-17A | SELP+ CEC | 0.484 | 0.094 | 1.836 |
| IL-8 | CX3CL1+ CEC | 0.614 | **0.026** | 2.580 |
| IL-18 | CX3CL1+ CEC | 0.591 | **0.033** | 2.432 |
| ***Convalescent COVID without CV risks*** | | | | |
| EGF | CEC | 0.660 | **0.020** | 2.778 |
| LIF | CEC | 0.800 | **0.002** | 4.210 |
| PDGF-BB | SELP+ CEC | 0.672 | **0.017** | 2.869 |
| PIGF-1 | CX3CL1+ CEC | 0.635 | **0.026** | 2.600 |
| IL-1RA | CEC | 0.546 | 0.066 | 2.062 |
| IP-10 | CEC | 0.706 | **0.010** | 3.152 |
| CXCL12 | CEC | 0.631 | **0.028** | 2.571 |
| RANTES | SELP+ CEC | 0.456 | 0.136 | 1.622 |
| IL-5 | CEC | 0.734 | **0.007** | 3.418 |
| IL-7 | SELP+ CEC | 0.753 | **0.005** | 3.623 |
| IL-18 | SELP+ CEC | 0.547 | 0.066 | 2.066 |
| IL-4 | CX3CL1+ CEC | 0.548 | 0.065 | 2.070 |

*Bold *p* values indicate statistical significance at alpha level 0.05 with non-parametric partial correlation coefficients calculated based on Spearman's rank correlation. n= 14 for Convalescent COVID with CV risks and n= 13 for Convalescent COVID without CV risks.

### Supplemental Table 3

Key findings from published studies on immunophenotyping of COVID-19 and convalescent patients.

| | Samples | Cell types | Approach | Lymphopenia phenotype | Terminal differentiation or exhaustion phenotype | Activation phenotype | Other notable phenotypes | References |
|---|---|---|---|---|---|---|---|---|
| 1 | 12 healthy, 7 recovered, 7 mild disease, 27 severe disease | PBMCs | High- dimensional flow cytometry | yes (severe disease, CD8+, CD4+, NK) | yes | yes, (heterogenous among severe patients) | Higher proportion of neutrophil and eosinophil populations in severe COVID-19+.<br>No lymphopenia in recovered patients.<br>Increased neutrophil to lymphocyte ration in severe COVID patients compared to mild or recovered patients.<br>Higher proportion of cytotoxic CD8+ T cells in severe COVID-19 individuals . | Kuri- Cervantes, L. et al. **Comprehensive mapping of immune perturbations associated with severe COVID-19. Sci**. Immunol.5, eabd7114 (2020) |
| 2 | 6 healthy, 3 non-ventilated, 4 with ARDS | PBMCs | scRNA- seq | yes( ARDS patients, NK) | no (T cell)/ Yes (NK) | yes (some ARDS patient) | Large heterogeneity of differentially expressed genes in COVID CD4+ T, CD8+ T and NK cell subset when compared to healthy donor, including of interferon- stimulated genes.<br>No substantial expression of pro-inflammatory cytokine genes by monocytes, T or NK cells. | Wilk, A. J. et al. **A single- cell atlas of the peripheral immune response in patients with severe COVID-19**. Nat. Med.26, 1070–1076 (2020). |
| 3 | 4 healthy , 5 severe influenza , 5 mild and 5 severe COVID disease | PBMCs | scRNA- seq | yes (severe disease, CD8+, CD4+, NK) | no reported | no reported | TNF/IL-1β inflammatory signature observed in all PBMCs found in COVID samples. | Lee, J.S. et al. 2020 **Immunophenotyping of COVID-19 and influenza highlights the role of type I interferons in development of severe COVID-19**. Sci. Immunol.5, eabd1554 (2020) |
| 5 | 60 healthy,<br>36 recovered,<br>125 hospitalized patients (NIH ordinal score 2–5) | PBMCs | High- dimensional flow cytometry | yes (severe disease, biais toward CD8+) | yes (heterogenous in COVID patients compared to healthy or recovered patients) | yes (heterogenous in COVID patients compared to healthy or recovered) | heterogeneity of T cell responses based on high- dimensional immune profiling, with three potential immune subtypes, all associated to mortality. | Mathew, D. et al. **Deep immune profiling of COVID-19 patients reveals distinct immunotypes with therapeutic implications**. Sciencehttps://doi.org/10.1126/science.abc8511 (2020). |
| 4 | 10 healthy, 10 ARDS patients | PBMCs | Flow cytometry | yes (ARDS patients, CD3+ T cells) | no reported | yes in ARDS samples | SARS-CoV-2–specific memory CD4+ and effector CD8+ T cells appear in blood of patients with ARDS 2 weeks post symptom onset.<br>SARS- CoV-2- specific T cells have a TH1 cell cytokine profile in ARDS patients.<br>Presence of cross-reactive lymphocyte in healthy samples possibly induced by seasonal coronaviruses. | Weiskopf, D. et al. **Phenotype and kinetics of SARS- CoV-2-specific T cells in COVID-19 patients with acute respiratory distress syndrome**. Sci. Immunol.5, eabd2071 (2020) |
| 6 | 20 healthy,<br>20 COVID convalescent | PBMCs | Flow cytometry | no reported | no reported | no reported | 100% of the CD4+ and 70% of the CD8+ T cells from convalescent COVID patients respond to SARS- CoV-2 epitopes, including S, M and N proteins and other ORFs.<br>T cell reactivity to SARS-CoV-2 is also detected in healthy donors possibly due to potential cross- reactivity to other common cold coronaviruses. | Grifoni, A. et al. **Targets of T cell responses to SARS- CoV-2 coronavirus in humans with COVID-19 disease and unexposed individuals**. Cell181, 1489–1501.e15 (2020). |
| 7 | 16 healthy, 28 mild recovered, 14 severe recovered | PBMCs | Flow cytometry | no reported | no reported | no reported | T cells from convalescent patients with mild or severe disease respond to SARS-CoV-2 epitopes<br>higher breadth and magnitude of T cell responses in severe as compared with mild convalescent;<br>No significant differences in the cytotoxic potential in patients with mild and severe disease;<br>Little Cytotoxic CD4+ T cells observed in recovered patient.<br>higher proportions of SARS-CoV-2-specific CD8+ T cells in mild convalescent | Peng, Y. et al. **Broad and strong memory CD4 and CD8 T cells induced by SARS-CoV-2 in UK convalescent COVID-19 patients**. Nat Immunol (2020). https://doi.org/10.1038/s41590-020-0782-6 |
| 8 | 15 healthy, 15 recovered, 24 acute COVID (mild to severe disease) | PBMCs | no reported | no reported | no reported | no reported | Coordinated responses of SARS-CoV-2-specific CD4+ and CD8+ T cell and neutralizing antibodies were associated to milder disease, with prominent roles for SARS-CoV-2-specific CD4+ T cells and to some extend CD8+ while neutralizing antibodies alone are not associated to protection.<br>CXCL10 may be a biomarker in acute COVID-19 of impaired T cell responses<br>ageing and depletion of naive T cells are associated to a decreased coordinated adaptive immune response, resulting in poor disease outcomes.<br>SARS-CoV-2-specific IFNg+ CD8+ T cells predominantly expressed granzyme B, with detectable TNFa and absence of IL-10, with a functional profile comparable to that of CMV-specific CD8 T cells | Rydyznski Moderbacher, C. et al. **Antigen-specific adaptive immunity to SARS-CoV-2 in acute COVID-19 and associations with age and disease severity**. Cell (2020). https://doi.org/10.1016/j.cell.2020.09.038 |
